# Evidence-Based Assessment Benchmarks and Bipolar Classification in ABCD Youth

**DOI:** 10.64898/2026.05.06.26352558

**Authors:** Eric A. Youngstrom, Amanda J. Thompson, Yinuo Liu, Marissa B. McClellan, Cristian Alcaino, Phoebe A. Rodda, Donna Ruch

## Abstract

**Objective:** To test whether two brief mania measures, the Parent General Behavior Inventory-10 Mania form (PGBI-10M) and 7-Up, retain useful psychometric properties in a large population cohort, and to evaluate whether the PGBI-10M can identify Kiddie Schedule for Affective Disorders and Schizophrenia (KSADS)-defined bipolar spectrum disorders in that setting.

**Method:** Analyses used 11,000+ youths across late childhood and early adolescence from the Adolescent Brain Cognitive Development (ABCD) Study. For both PGBI-10M and 7-Up, we estimated descriptive statistics, internal consistency, confirmatory factor models, graded response models, and measurement-based care benchmarks (minimally important difference, reliable change, and clinical cutpoints). For the PGBI-10M, receiver operating characteristic (ROC) analyses estimated concurrent classification accuracy for bipolar diagnoses at baseline and 2-year follow-up and compared area under the curve (AUC) values with prior outpatient and community mental health samples.

**Results:** Scores were lower than in clinical samples, but both measures remained psychometrically sound. The PGBI-10M showed alpha=.87–.88 and omega=.88; the 7-Up showed alpha=.78 and omega=.79. Longitudinal analyses indicated threshold differences across waves, likely reflecting caregiver recalibration and developmental changes, with modest impact on estimates. ABCD-based benchmarks supported meaningful and reliable change. The PGBI- 10M discriminated bipolar cases (AUC=0.68 baseline; 0.77 follow-up), though performance was lower than in clinical samples. Positive predictive values were low in this population.

**Conclusion:** The PGBI-10M and 7-Up retain sound psychometric properties in a large general population cohort, supporting population-referenced benchmarks for symptom elevation most suited to single-timepoint referral triage and indication-based assessment rather than universal screening.

## Introduction

Routine use of brief, standardized measures to monitor symptoms and functioning can improve detection, facilitate timely treatment adjustment, and is associated with better outcomes in both adult and youth settings.^1,2^ Yet most pediatric measurement-based care (MBC) efforts have focused on depression, anxiety, and general distress, leaving a clinically vital gap in tools for tracking hypomanic, manic, and mixed symptoms in youths. Bipolar spectrum disorders in youth are associated with substantial functional impairment, risk for self-harm, and long-term mood instability,^3–5^ yet diagnosis is often delayed by years.^6,7^ Early and accurate identification remains a priority in child and adolescent mental health care.^8^

Brief rating scales are central to evidence-based assessment, and carefully selected measures can improve identification of bipolar spectrum disorders when integrated with clinical interviewing and other assessment data.^9,10^ Among available options, the Parent General Behavior Inventory-10 Mania form (PGBI-10M) has particularly strong support. Derived from the longer General Behavior Inventory,^11^ it is a 10-item, freely available parent-report scale assessing hypomanic, manic, and mixed mood symptoms in youths.^12^ In academic outpatient and community mental health settings, the PGBI-10M has shown excellent internal consistency and strong discriminative validity for KSADS-defined bipolar spectrum disorders,^9,10^ and meta-analyses put it in the top tier of scales for pediatric bipolar assessment.^13,14^ The youth-report 7- Up scale is another promising brief measure, designed to capture elevated mood, activation, and decreased need for sleep from the young person’s perspective.^15^ Prior work suggests it may help monitor mania-related symptoms in adolescents and young adults.^15,16^ Together, these measures are attractive candidates for brief, population-referenced assessment; shorter-recall versions of these scale families have also been used for repeated MBC monitoring in treatment contexts.^17,18^

The evidence base has an important limitation: most research on pediatric mania scales has been conducted in help-seeking or clinically enriched samples. Test performance depends heavily on sampling frame. In specialty settings, youths with bipolar spectrum disorders tend to present with more severe mood symptoms, greater impairment, and richer comorbidity, widening the separation between bipolar and non-bipolar groups on mania-related scales.^19,20^ In general population samples, by contrast, symptom levels are lower and more restricted,^21^ the bipolar base rate is low,^22–24^ and many youths with elevated scores may instead have externalizing problems, trauma-related symptoms, or other conditions that complicate differential diagnosis.^25,26^ Strong performance in clinical samples does not guarantee comparable accuracy in community cohorts.

The gap is not only about classification. For MBC, clinicians also need benchmarks for interpreting symptom change over time. Many bipolar rating scales were developed for diagnostic assessment rather than repeated monitoring, and relatively few have well-developed interpretive anchors such as minimally important differences,^27^ reliable change indices, and clinically significant change thresholds in general population or mixed-severity samples.^28,29^ Without such anchors, it is difficult to determine when score changes are large enough to matter clinically, or when symptom levels are substantially elevated relative to age peers. These two forms of benchmark serve different purposes: change-oriented indices such as the MID and reliable change index quantify how much a score must shift over time to be considered clinically meaningful, whereas nomothetic milestones—population-referenced percentile or standard-deviation cutoffs—benchmark a single score against age-peer norms without requiring repeated measurement. The need for normative reference points is especially acute in child and adolescent mood assessment, where many relevant behaviors lie on a continuum with ordinary developmental variation.^30,31^

The Adolescent Brain Cognitive Development (ABCD) Study provides a rare opportunity to address these questions in a single population-based cohort. ABCD is a large, multisite longitudinal study of U.S. youth with extensive demographic and mental health assessment across late childhood and early adolescence.^32,33^ Crucially, ABCD includes the PGBI-10M and the youth-report 7-Up in a large, demographically diverse, nonreferred sample, and it also includes structured KSADS-based bipolar spectrum diagnoses at key years.^32–34^ This design makes it possible to examine two complementary questions in the same setting: whether the PGBI-10M and 7-Up retain useful psychometric properties outside specialty care and support population-referenced interpretive benchmarks, and whether the PGBI-10M can discriminate youths with KSADS-defined bipolar spectrum disorders from those without such diagnoses in a large community cohort.

The present study therefore had three aims. First, we evaluated the psychometric performance of the PGBI-10M and 7-Up in ABCD, including score distributions, internal consistency, dimensionality, item response characteristics, and population-referenced interpretive benchmarks. Based on prior work, we expected both measures to show adequate to strong reliability and predominantly unidimensional structure,^10,16^ with some attenuation relative to clinical samples because of restricted range. A second, MBC-focused aim was to derive population-referenced interpretive benchmarks—minimally important differences (MID), reliable change indices (RCI), and clinical significance thresholds—that clinicians could use to judge whether a given symptom level is elevated relative to age peers and, when repeated assessment is available, whether an observed change exceeds measurement error. Third, we evaluated the PGBI-10M’s discriminative validity. We expected it to be statistically significant, but to perform less strongly in ABCD than in clinically enriched settings because of lower prevalence, milder average severity, and greater overlap between bipolar and non-bipolar presentations.^19–21^

By combining psychometric benchmarking with diagnostic classification in the same cohort, this study aims to clarify the appropriate role of brief pediatric mania scales in practice. The goal is not to treat a single score as a diagnostic verdict, but to determine when these measures are most useful: as tools for monitoring manic and mixed symptoms over time, for identifying clinically meaningful change, and for informing targeted, multi-informant, algorithm-assisted assessment when bipolar disorder is a plausible concern.

## Method

### Participants and Procedure

This study analyzed data from the Adolescent Brain Cognitive Development (ABCD) Study, an ongoing multisite longitudinal cohort of youths recruited at ages 9 to 10 years from schools and communities across 21 U.S. sites. ABCD was designed to approximate the demographic composition of U.S. youth in this age range and includes extensive behavioral and mental health assessment across development. Parents or legal guardians provided written informed consent, and youths provided assent. Study procedures were approved by institutional review boards at participating sites.

We used data from baseline and the first three annual follow-up assessments of the ABCD study (release 5.1 doi.org/10.15154/z563-zd24). Psychometric and measurement-based care (MBC) analyses used years in which the Parent General Behavior Inventory-10 Mania form (PGBI-10M) and youth-report 7-Up were administered: baseline and Year 2 for the PGBI-10M, and Year 1 and Year 3 for the 7-Up. Classification analyses used baseline and Year 2 follow-up, when both PGBI-10M and concurrent KSADS bipolar spectrum diagnoses were available. We also used data from a study in outpatient mental health clinics^10^ for direct comparison of the classification accuracy of the PGBI-10M. The pooled outpatient comparison sample (*N*=1,355) exceeds the combined subsample total reported in the original source (*N*=1,147) because that publication required complete data on the Achenbach measures for its convergent-validity analyses, whereas the present analyses use only PGBI-10M/7-Up items and diagnostic status.

Psychometric analyses included participants with sufficient item-level data to calculate a valid total score. Classification analyses required both a valid PGBI-10M total score and diagnostic data sufficient to determine presence or absence of a bipolar spectrum diagnosis at the same year. Participants could contribute data at some years and not others, depending on attrition and missingness.

### Measures

#### Parent General Behavior Inventory-10 Mania Form

The PGBI-10M is a 10-item parent-report measure of hypomanic, manic, and mixed mood symptoms in youths^12^ carved from the General Behavior Inventory (GBI) to maximize discrimination between bipolar spectrum disorders and other pediatric conditions.^10^ Caregivers rate each item using a 0-3 response format, and the total score sums them, with higher values indicating more frequent or severe manic or mixed symptoms.

In ABCD, primary caregivers completed the PGBI-10M using “past year” as the reference period. Psychometric analyses used scores at baseline and Year 2. When 20% or fewer items were missing, totals were prorated; when >20% were missing, totals were coded missing. Factor and item response theory analyses used FIML estimation with item level data.

#### Youth Report 7-Up

The 7-Up was designed as a self-report measure of elevated mood and behavioral activation, including items reflecting increased energy, decreased need for sleep, and related experiences, also carved from the GBI. Whereas the 10M was designed to maximize discriminative validity, the 7-Up item selection focused on internal consistency and factor validity.^15^ Item scores are summed to produce a total score, with higher scores indicating greater manic or activation tendencies. Prior work supports the psychometric utility of related self-report short forms for mood symptoms in adolescents and young adults.^15,16^

In ABCD, the 7-Up was administered to youths at follow-up visits, also using “past year” as reference period. Total scores were used in psychometric and MBC analyses at Year 1 and Year 3. Missing-data rules paralleled those for the PGBI-10M. The 7-Up was not included in concurrent classification analyses because it was not consistently administered at the same time as the KSADS interview.

#### KSADS Bipolar Spectrum Diagnoses

Bipolar spectrum diagnoses were derived from computerized KSADS assessments administered within ABCD.^34^ A bipolar-positive outcome was defined as any bipolar spectrum diagnosis as reported by parent or youth at the target year, including bipolar I, bipolar II, cyclothymic disorder, or other specified/unspecified bipolar and related disorder, based on ABCD diagnostic summary variables corresponding to ICD-10 coding. Youths without any bipolar spectrum diagnosis at that year were coded bipolar-negative. Primary classification analyses focused on concurrent discrimination: PGBI-10M scores at baseline and Year 2 predicting KSADS bipolar spectrum diagnoses at the same year.

### Statistical Analysis

Analyses addressed three aims: evaluation of psychometric performance for the PGBI-10M and 7-Up; derivation of population-referenced MBC benchmarks (MID, RCI, clinical significance thresholds); and evaluation of concurrent classification accuracy for the PGBI-10M.

We first summarized score distributions and missingness for each scale and year. Psychometric analyses followed the analytic protocol in prior work and best practices in scale development and validation.^35^ Cronbach’s alpha, McDonald’s omega, and mean inter-item correlations evaluated internal consistency using the *psych* package (v2.6.5) in R.^36^ Single-factor confirmatory factor models were fit separately for each scale with estimators appropriate for ordered categorical indicators using *lavaan* v0.7-2.^37^ Comparative fit index, Tucker-Lewis index, root mean squared error of approximation, and standardized root mean square residual evaluated fit.

To characterize precision across the latent severity continuum, we fit Samejima graded response models to item-level data for each scale and year using *mirt* v1.4.6.1.^38^ These models derived item and test information functions and converted test information to conditional reliability, identifying the symptom range over which each scale achieved adequate reliability for individual-level decision making. To evaluate longitudinal measurement equivalence, we conducted measurement invariance analyses using multigroup graded response models. For each instrument, we compared configural, metric, and scalar models across years, testing whether item discrimination parameters and response thresholds were stable across measurement occasions.

Sequential likelihood ratio tests with FDR correction identified items showing differential functioning across years. Partial invariance models were estimated when full scalar invariance was not supported. Longitudinal stability of IRT-derived factor scores was evaluated against raw score stability using Pearson correlations in matched longitudinal subsamples.

To support MBC applications, we derived minimally important differences, reliable change indices, and clinical significance thresholds. Minimally important differences were estimated using a distribution-based approach anchored to approximately one-half standard deviation of the observed score distribution.^27^ Reliable change indices were calculated following Jacobson and Truax,^28^ using observed score standard deviations and omega-based reliability estimates to derive raw-score changes corresponding to conventional confidence thresholds. Clinical significance thresholds were defined using score values approximately 1.95 standard deviations above the ABCD mean, supplemented by percentile-based benchmarks for clinical translation.

Concurrent classification accuracy of the PGBI-10M was evaluated with receiver operating characteristic curves at baseline and Year 2, using PGBI-10M total score as the index test and KSADS bipolar spectrum diagnosis as the criterion. Area under the curve (AUC) was estimated nonparametrically, with standard errors and 95% confidence intervals using *pROC* v1.19.0.1.^39^ ABCD AUCs were compared with published values from prior academic outpatient and community mental health samples using Hanley and McNeil’s^40^ method for independent ROC curves. Exploratory subgroup analyses used logistic regression to test whether base rates or score-outcome associations varied across sex and race/ethnicity contrasts.

## Results

### Sample Characteristics and Score Distributions

Table 1 summarizes sample characteristics by year. We included all available data from male and female youth participating in ABCD, ranging from 11,864 youths at baseline to 10,332 at Year 3. The sample was roughly evenly split by sex, and just over half of participants were identified as White at each year. Mean age was 9.92 years (*SD*=0.62) at baseline and 12.03 years (*SD*=0.67) at Year 2. Mood disorder diagnoses were uncommon, with bipolar spectrum diagnoses identified in 2.0% of youths at baseline and 1.2% at Year 2.

**Table 1.** Demographics and clinical characteristics by ABCD Wave, and Outpatient Mental Health comparison sample

|  | *Year 0* | *Year 1* | *Year 2* | *Year 3* | *Outpatient MH* |
| --- | --- | --- | --- | --- | --- |
| *Youth Demographics* |  |  |  |  |  |
| *N* | 11,864 | 11,216 | 10,969 | 10,332 | 1355 |
| Female, % (*n*) | 47.8% (5,676) | 47.7% (5,348) | 47.7% (5,216) | 47.7% (4,908) | 39.0% (547) |
| Age, *M* (*SD*) | 9.92 (0.62) | 10.93 (0.64) | 12.03 (0.67) | 12.92 (0.65) | 11.05 (3.40) |
| White, % (*n*) | 52.0% (6172) | 53.3% (5984) | 53.4% (5860) | 54.1% (5591) | 45.5% (625) |
| Black, % (*n*) | 15.0% (1784) | 14.2% (1597) | 14.2% (1561) | 13.2% (1361) | 46.9% (647) |
| Hispanic, % (*n*) | 20.3% (2410) | 19.8% (2221) | 19.8% (2164) | 19.8% (2075) | 2.4% (33) |
| Other, % (*n*) | 12.6% (1499) | 12.6% (1415) | 12.6% (1386) | 12.6% (1307) | 4.8% (66) |
| *Clinical Characteristics* |  |  |  |  |  |
| **Any mood disorder diagnosis, % (*n*)** | 3.3% (385) | -- | 2.6% (288) | -- | 56.0% (766) |
| **Bipolar spectrum diagnosis, % (*n*)** | 2.0% (235) | -- | 1.2% (129) | -- | 30.1% (414) |

**Note.** ABCD = Adolescent Brain Cognitive Development study. “—” indicates structural missingness, where the variable was not available in ABCD. ABCD samples include all available data for male and female youth within each assessment period. Outpatient mental health (MH) sample is secondary analysis of NIH R01 MH066647 and Stanley Medical Research Institute pooled data. The pooled Outpatient MH *N* (1,355) is larger than the combined subsample total in the original source (*N*=1,147) because that study's analyses required complete Achenbach data, which is not a requirement here (see Methods).

Table 2 presents descriptive statistics for the PGBI-10M and 7-Up. Mean scores were low across years, consistent with the nonreferred nature of the ABCD cohort. PGBI-10M, *M*=1.30 (*SD*=2.77) at baseline and 1.11 (*SD*=2.57) at Year 2. For the 7-Up, means were 2.25 (*SD*=2.87) at Year 1 and 1.81 (*SD*=2.44) at Year 3. Observed score ranges extended well into the upper end of each scale despite the low means, indicating that a minority of youths showed marked elevations in manic or activation symptoms. Overall, these distributions were notably lower and more restricted than would be expected in clinically ascertained samples.

**Table 2.**
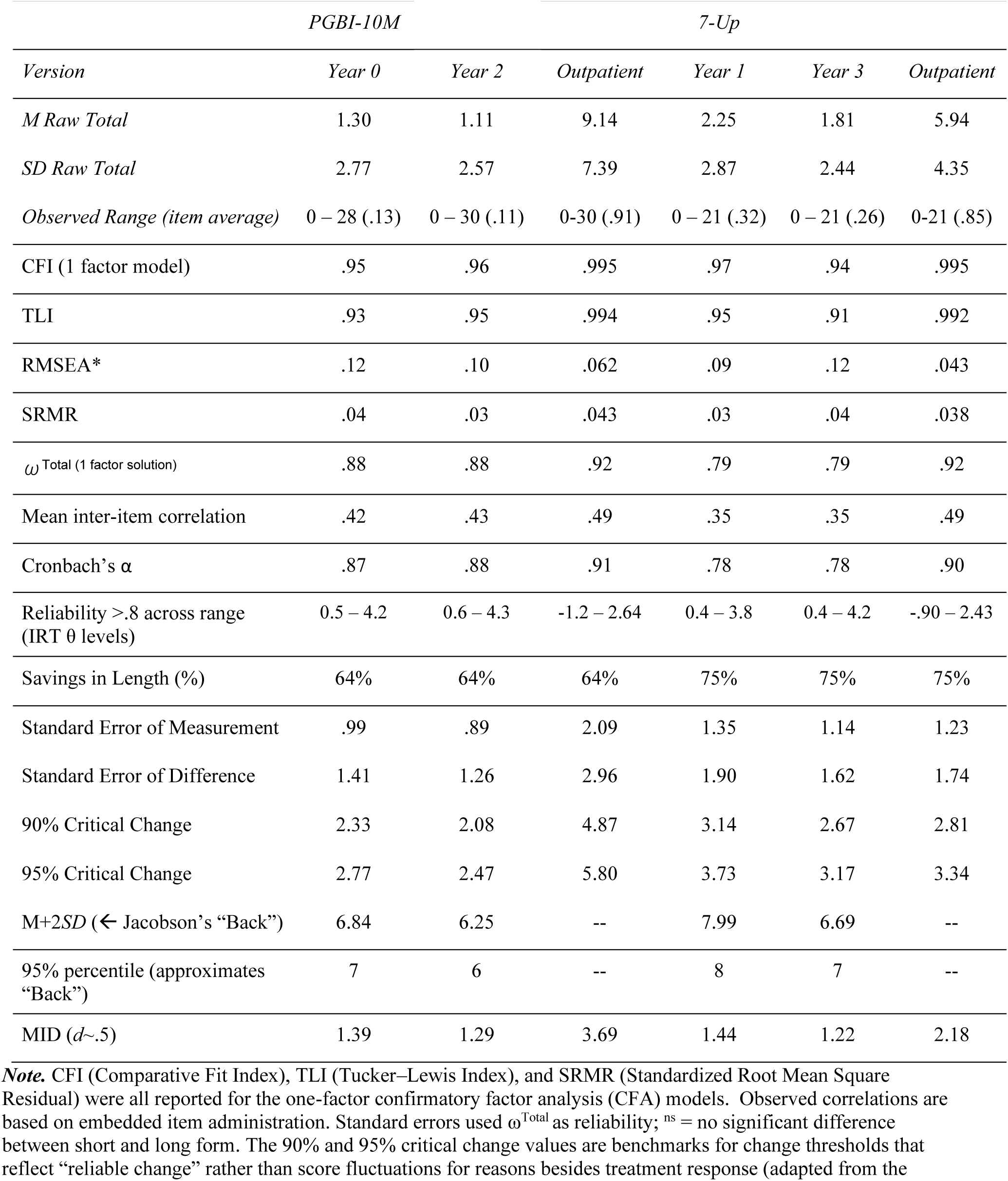
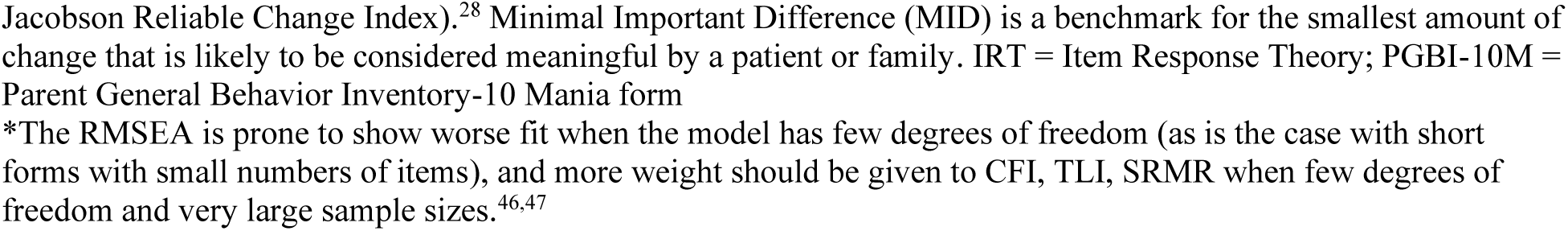
Descriptive statistics, internal consistency reliability, and clinical significance benchmarks for PGBI-10M and 7-Up by Wave and with Outpatient Mental Health Comparator^44,45^

|  | PGBI-10M |  |  | 7-Up |  |  |
| --- | --- | --- | --- | --- | --- | --- |
| Version | Year 0 | Year 2 | Outpatient | Year 1 | Year 3 | Outpatient |
| *M Raw Total* | 1.30 | 1.11 | 9.14 | 2.25 | 1.81 | 5.94 |
| *SD Raw Total* | 2.77 | 2.57 | 7.39 | 2.87 | 2.44 | 4.35 |
| *Observed Range (item average)* | 0 – 28 (.13) | 0 – 30 (.11) | 0-30 (.91) | 0 – 21 (.32) | 0 – 21 (.26) | 0-21 (.85) |
| CFI (1 factor model) | .95 | .96 | .995 | .97 | .94 | .995 |
| TLI | .93 | .95 | .994 | .95 | .91 | .992 |
| RMSEA* | .12 | .10 | .062 | .09 | .12 | .043 |
| SRMR | .04 | .03 | .043 | .03 | .04 | .038 |
| $\omega$ Total (1 factor solution) | .88 | .88 | .92 | .79 | .79 | .92 |
| Mean inter-item correlation | .42 | .43 | .49 | .35 | .35 | .49 |
| Cronbach's $\alpha$ | .87 | .88 | .91 | .78 | .78 | .90 |
| Reliability >.8 across range (IRT $\theta$ levels) | 0.5 – 4.2 | 0.6 – 4.3 | -1.2 – 2.64 | 0.4 – 3.8 | 0.4 – 4.2 | -.90 – 2.43 |
| Savings in Length (%) | 64% | 64% | 64% | 75% | 75% | 75% |
| Standard Error of Measurement | .99 | .89 | 2.09 | 1.35 | 1.14 | 1.23 |
| Standard Error of Difference | 1.41 | 1.26 | 2.96 | 1.90 | 1.62 | 1.74 |
| 90% Critical Change | 2.33 | 2.08 | 4.87 | 3.14 | 2.67 | 2.81 |
| 95% Critical Change | 2.77 | 2.47 | 5.80 | 3.73 | 3.17 | 3.34 |
| M+2SD (← Jacobson's "Back") | 6.84 | 6.25 | -- | 7.99 | 6.69 | -- |
| 95% percentile (approximates "Back") | 7 | 6 | -- | 8 | 7 | -- |
| MID ($d \sim .5$) | 1.39 | 1.29 | 3.69 | 1.44 | 1.22 | 2.18 |

**Note.** CFI (Comparative Fit Index), TLI (Tucker–Lewis Index), and SRMR (Standardized Root Mean Square Residual) were all reported for the one-factor confirmatory factor analysis (CFA) models. Observed correlations are based on embedded item administration. Standard errors used $\omega^{\text{Total}}$ as reliability; <sup>ns</sup> = no significant difference between short and long form. The 90% and 95% critical change values are benchmarks for change thresholds that reflect “reliable change” rather than score fluctuations for reasons besides treatment response (adapted from the
Jacobson Reliable Change Index).<sup>28</sup> Minimal Important Difference (MID) is a benchmark for the smallest amount of change that is likely to be considered meaningful by a patient or family. IRT = Item Response Theory; PGBI-10M = Parent General Behavior Inventory-10 Mania form
\*The RMSEA is prone to show worse fit when the model has few degrees of freedom (as is the case with short forms with small numbers of items), and more weight should be given to CFI, TLI, SRMR when few degrees of freedom and very large sample sizes.<sup>46,47</sup>

### Reliability and Dimensionality of the PGBI-10M and 7-Up

Both measures showed acceptable to strong internal consistency in ABCD, and even higher in the outpatient mental health sample (see Table 2). Both scales retained coherent internal structure in a general population cohort, although reliability was somewhat stronger for the caregiver-report PGBI-10M than for the youth-report 7-Up. The latent-variable results were also consistent with useful unidimensional measurement. Single-factor models fit well for both scales, with fit indices in ranges conventionally interpreted as good, supporting treatment of the PGBI-10M and 7-Up total scores as primarily indexing a common manic or activation dimension. Figure 1 displays the item option characteristic curves for both measures.

**Figure 1.**
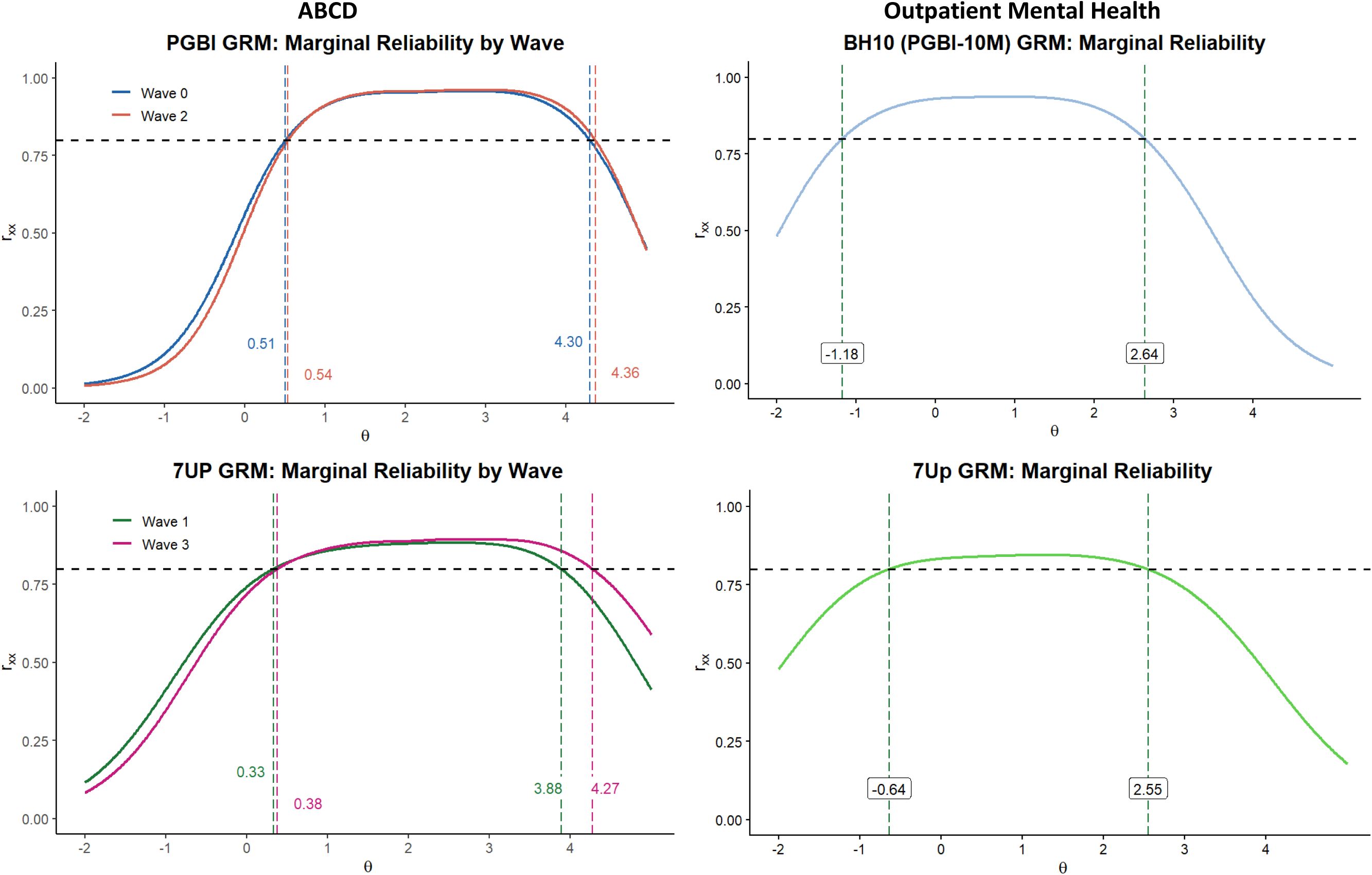
Conditional Reliability of the PGBI-10M and 7-Up Across the Manic Symptom Continuum: Both Instruments Achieve Strong Reliability (*Rxx*) in the Clinically Elevated Range ***Note.*** Dashed horizontal line indicates reliability = .80. Both instruments exceed this threshold across the clinically elevated symptom range (θ > 0.5) at both waves, despite partial measurement non-equivalence across occasions. GRM = Graded Response Model; PGBI-10M = Parent General Behavior Inventory-10 Mania form

**Figure 2.**
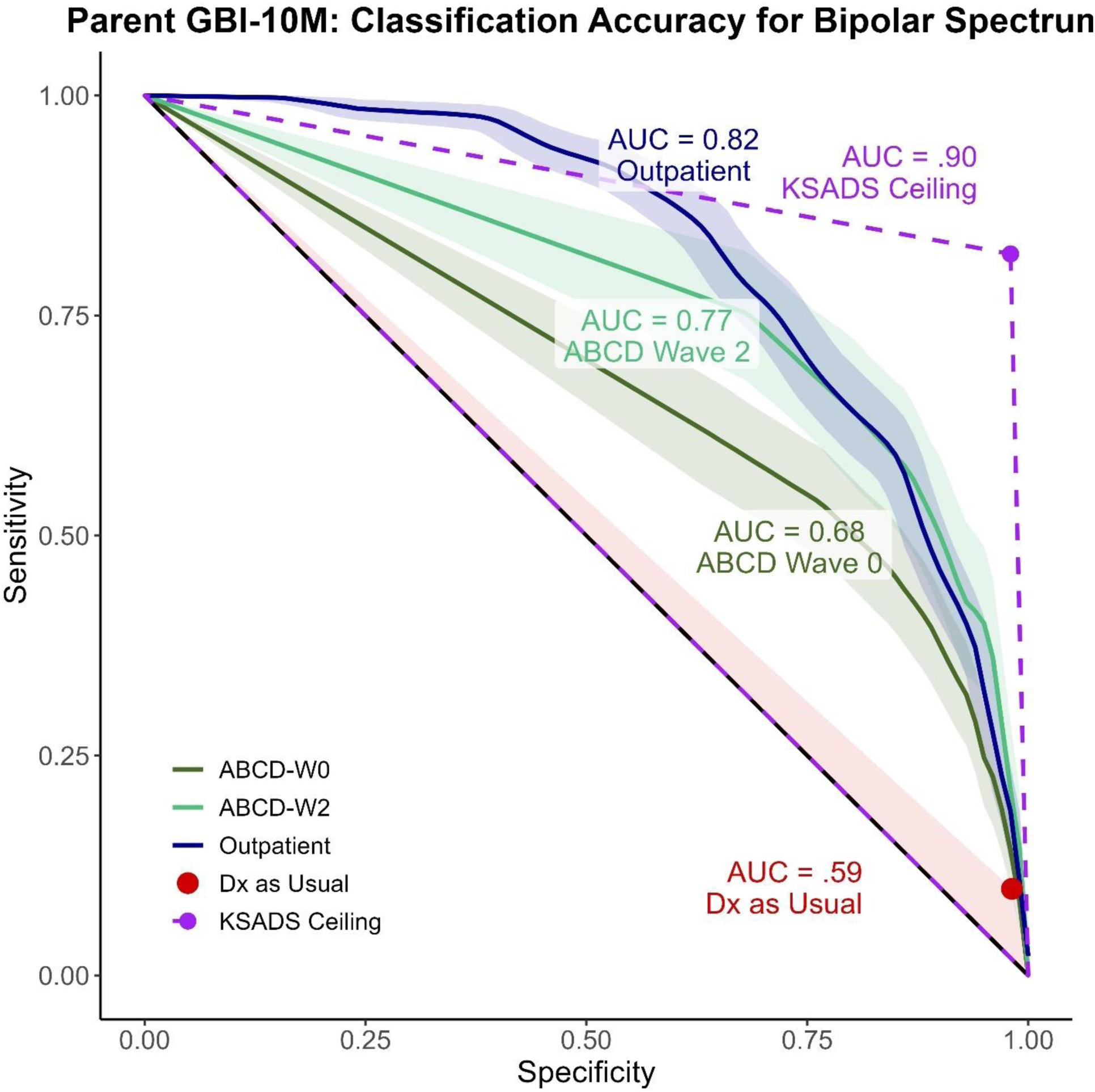
Areas under the curve for the PGBI-10M and chance performance, diagnosis as usual, and upper bound based on accuracy of the KSADS ***Note.*** Diagnosis (Dx) as Usual estimates accuracy of diagnosis as usual using published estimates^48,49^ combined with a base rate of 4%.^24^ The KSADS ceiling is set by the kappa of the KSADS (∼.90) and the base rate, using the formula from Kraemer.^21^ The shading around the curves for ABCD Wave 0, Wave 2, and Outpatient curves indicate the standard errors. Outpatient data are a secondary analysis pooling an academic outpatient and community mental health center.^44,45^ The accuracy of the PGBI-10M in the outpatient sample is in the top tier of mania scales based on recent meta-analysis,^13^ whereas the Wave 0 would be considered fair, and the Wave 2 falls in between. ABCD = Adolescent Brain Cognitive Development study; AUC = Area Under the Curve; KSADS = Kiddie Schedule for Affective Disorders and Schizophrenia; PGBI-10M = Parent General Behavior Inventory-10 Mania form.

### Item Response Theory Findings and MBC Benchmarks

Item response theory analyses indicated that both scales provided clinically useful information across elevated portions of the latent severity continuum. For the PGBI-10M, conditional reliability exceeded .80 from theta levels of approximately 0.5 to 4.2 at baseline and 0.6 to 4.3 at Year 2. For the 7-Up, reliability exceeded .80 from approximately 0.4 to 3.8 at Year 1 and 0.4 to 4.2 at Year 3. Thus, both instruments provided their strongest measurement precision in the range most relevant to identifying elevated manic-spectrum symptoms, despite restricted distributions in this nonclinical sample. Figure 1 shows the reliability curves for both scales at both years; supplemental figure 1 shows the option characteristic curves.

Table 2 also provides benchmarks intended to support measurement-based care. Standard errors of measurement were 0.99 and 0.89 for the PGBI-10M at baseline and Year 2, respectively, and 1.35 and 1.14 for the 7-Up at Year 1 and Year 3. Corresponding 95% critical change values were 2.77 and 2.47 for the PGBI-10M and 3.73 and 3.17 for the 7-Up. In practice, raw-score changes of roughly 3 points on either measure would often be needed before a clinician could be confident that observed change exceeded measurement error, with somewhat smaller thresholds for the PGBI-10M at Year 2 and somewhat larger thresholds for the 7-Up. MID estimates were smaller, ranging from 1.22 to 1.44 across scales and years, suggesting that clinically meaningful shifts may occur before changes become large enough to satisfy strict reliable-change criteria.

Population-referenced clinical significance thresholds were also modest in raw-score units because of the low means in ABCD. Mean +2 standard deviation values ranged from 6.25 to 6.84 for the PGBI-10M and from 6.69 to 7.99 for the 7-Up, with empirical 95th percentiles closely aligned at 6 to 8 points across years. Taken together, these benchmarks indicate that scores well below the maximum possible range may still represent substantial elevation relative to age peers in a general population sample.

### Longitudinal Measurement Equivalence

For the PGBI-10M, metric invariance (equal item discriminations across years) was supported with one exception, elated mood endorsed by others (Item #2), FDR *p*=.028. Scalar invariance was not fully supported: six of ten items showed threshold DIF (FDR *p*<.05). Invariant items were increased energy and elation (#4), decreased need for sleep (#6), and shifts in activity (#7) or mood and activity (#3) from high to low. Supplemental Table 1 reports item-level slopes and thresholds.

For the 7-Up, neither metric nor scalar invariance was fully supported; all six non-anchor items showed significant threshold DIF (all adjusted *p*≤ .012), with the largest shift in the excitement-seeking/novelty item (#2, *X²*=93.4). Partial invariance models accounting for these item-level differences yielded empirical reliability estimates of .58 (PGBI-10M) and .68 (7-Up). Longitudinal stability of IRT factor scores was *r*=.52 for the PGBI-10M (Years 0–2) and *r*=.33 for the 7-Up (Years 1–3), values nearly identical to raw score stability coefficients (.51 and .32, respectively), indicating that while measurement non-invariance was statistically significant, its practical impact on rank-order longitudinal estimates was modest (see Supplemental Table 2).

### Convergent and Discriminant Validity of the Scales

Table 3 presents the convergent and discriminant validity correlations for the PGBI-10M and 7-Up, with the coefficients grouped in descending order of expected correlation following the framework used in Youngstrom et al. (2020; 2021). Cohen’s *q* effect sizes quantified the difference between ABCD correlations versus in the outpatient mental health sample. The observed correlations showed high agreement in terms of rank order, and small to medium discrepancies with the outpatient sample, with the KSADS coefficients being among the most discrepant.

**Table 3.**
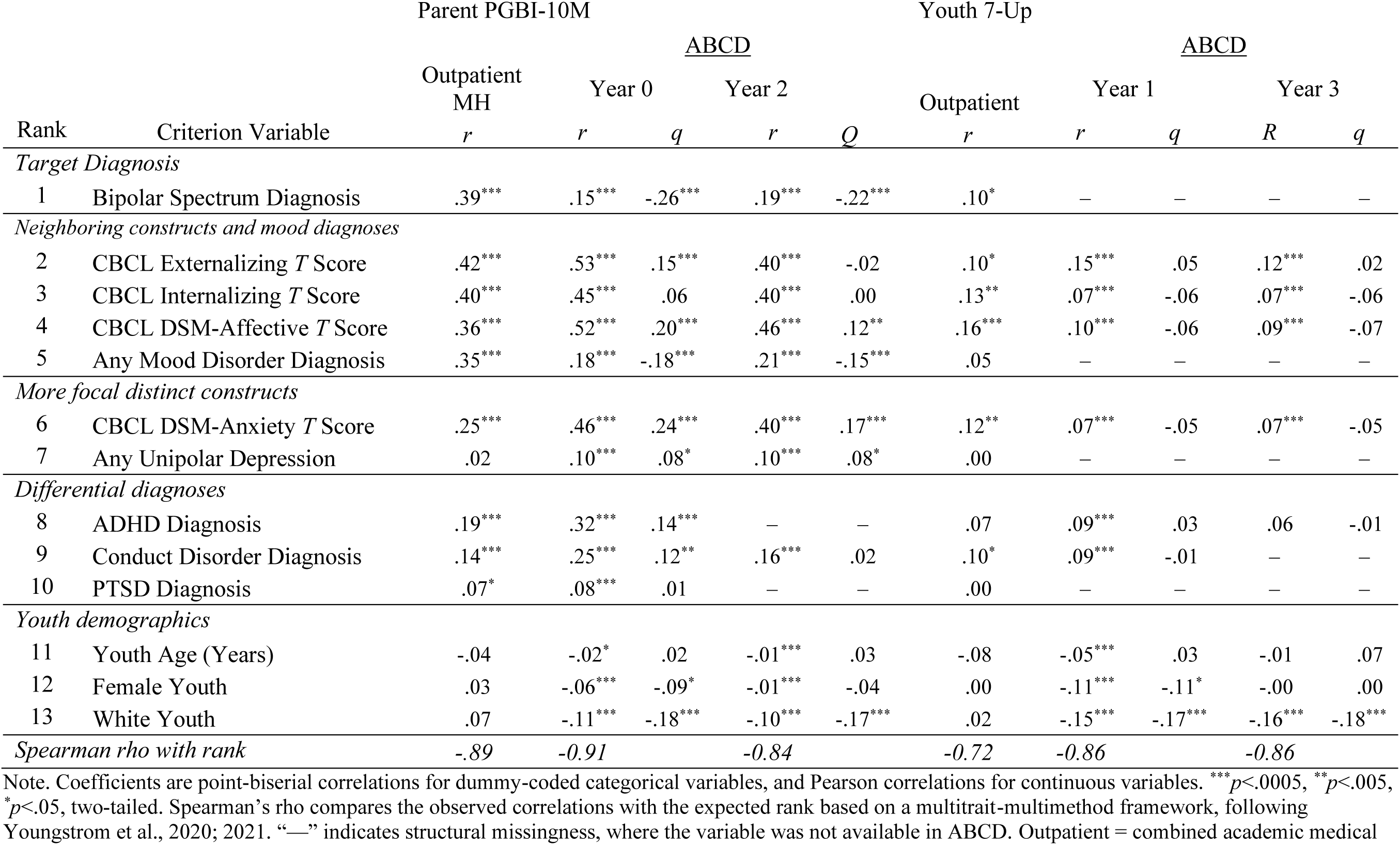
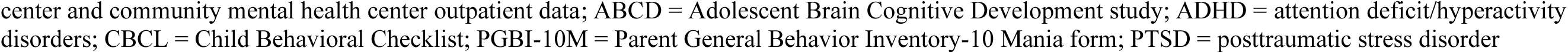
Convergent and discriminant validity of the parent-reported PGBI-10M and youth reported 7-Up, comparing performance in ABCD to corresponding coefficients with the same variables in outpatient mental health clinics.

### Classification Accuracy of the PGBI-10M for Bipolar Spectrum Diagnoses

The PGBI-10M significantly separated youths with KSADS-defined bipolar spectrum diagnoses from those without such diagnoses at both years. At baseline, the AUC=0.68 (*SE*=0.02, *p*<.0005). Year 2 classification accuracy improved to AUC=0.77 (*SE*=0.02, *p*<.0005). These findings indicate better-than-chance concurrent discrimination in the ABCD sample, with stronger separation at follow-up than at baseline.

Despite statistically significant AUCs, operating characteristics reflected the challenges of screening for a rare condition in a population sample. At cutpoints near 1.5 to 2 standard deviations above the ABCD mean, sensitivity was modest and specificity was high, but positive predictive values remained below 10% at both years, whereas negative predictive values exceeded 98%. This pattern indicates that elevated PGBI-10M scores identified youths at greater likelihood of bipolar-spectrum diagnoses, but that most youths with high scores in the general population still did not meet KSADS criteria for bipolar disorder.

### Exploratory Subgroup and Sensitivity Analyses

Exploratory subgroup analyses tested whether classification accuracy was moderated by sex, age, or race. Neither sex nor age showed significant main effects or interactions (albeit the age range is narrow in the ABCD data used). However, white versus nonwhite comparison revealed a small but significant interaction in Year 2, with white participants having a lower base rate of bipolar diagnoses, with odds of bipolar increasing more rapidly with each additional PGBI-10M point (OR 1.08/point, *p*=.013), with the odds crossing at a score of ∼8-10 points. The race interaction explained 0.6% of the variance and had a Cohen’s *d*=.04, where .2 would be small. Excluding participants with incomplete covariate data did not materially alter the overall pattern of findings. Thus, the principal results appeared robust: both scales remained psychometrically useful for measurement-based care applications, whereas the PGBI-10M showed statistically significant but attenuated concurrent classification accuracy for bipolar spectrum diagnoses in this general population sample.

### Comparisons With Prior Clinical Samples

Classification accuracy in ABCD was attenuated relative to previously reported results from clinically enriched samples. Prior academic outpatient studies reported AUC=0.84, and community mental health studies reported an AUC=0.78 for PGBI-10M prediction of KSADS bipolar diagnoses ^10,14^. Using Hanley and McNeil comparisons, the baseline ABCD AUC of 0.68 was significantly lower than both clinical benchmarks. The Year 2 ABCD AUC of 0.77 approached the community mental health estimate but remained lower than the academic outpatient value. This is consistent with the expectation that diagnostic discrimination is reduced in a nonreferred, low-base-rate cohort with milder average symptom severity and greater overlap between bipolar and nonbipolar presentations.

## Discussion

This study evaluated brief pediatric mania measures in a large, nonreferred cohort and yielded two main findings. First, both the PGBI-10M and the youth-report 7-Up kept useful psychometric properties in the ABCD Study. Internal consistency remained good, item response theory analyses showed clinically useful precision across elevated portions of the latent severity continuum, and the resulting benchmarks provided practical anchors for interpreting symptom severity and change over time in a nonclinical population. Second, the PGBI-10M significantly separated youths with KSADS-defined bipolar spectrum diagnoses from those without such diagnoses, but classification accuracy was attenuated relative to prior clinical samples, both in a comparison reported here and in meta-analyses that included other samples^13,14^. In ABCD, the PGBI-10M yielded AUCs of 0.68 at baseline and 0.77 at follow-up, indicating better-than-chance concurrent discrimination, yet positive predictive values remained low because bipolar spectrum disorders are generally uncommon in children in this age range. Taken together, these results suggest that brief mania scales can function well as tools for measurement-based assessment in general population youth, but they are not well suited for universal diagnostic screening.

The pattern fits what would be expected from evidence-based assessment principles. In clinically enriched samples, affected youths tend to have more symptoms, more impairment, and are more distinct from comparison groups, which tends to inflate diagnostic discrimination relative to community cohorts. In general population samples, symptom distributions are more mild and restricted,^19,20^ bipolar-spectrum conditions are rare, and many youths with elevated checklist scores will have other forms of psychopathology or subthreshold mood dysregulation rather than bipolar disorder. Under those conditions, a measure can remain psychometrically coherent while still showing weaker diagnostic separation.

The classification results point in the same direction. In a low-base-rate population, even a statistically informative test will often have poor positive predictive value when deployed indiscriminately. For that reason, the PGBI-10M is better understood as a tool for targeted assessment when bipolar disorder is already plausible because of presenting symptoms, course, family history, treatment-emergent activation, or other risk indicators. Evidence-based pediatric bipolar assessment has long argued for exactly this Bayesian approach: start with the base rate, add family and developmental history, then use questionnaires and interviews to revise probability rather than substitute for diagnosis. Clinically useful estimates of accuracy and thresholds are available^10,16^. The current results support that view. A high PGBI-10M score in a community sample should trigger closer evaluation, not function as a diagnostic verdict.

Psychometric usefulness for monitoring is not the same thing as stand-alone screening efficiency for a rare diagnosis. That distinction is clinically important because measurement-based care and screening are often conflated. Measurement-based care refers to repeated use of standardized symptom data to guide care decisions over time, and there is substantial support for it improving mental health treatment quality and outcomes. Currently, implementation remains incomplete in youth services, especially in routine community settings, where brief and accessible tools are still underused.^8^

The results pattern reflects a genuine distinction, not inconsistency: MID (*d*≈.5) estimates the smallest score change a patient is likely to notice, whereas RCI sets a stricter threshold beyond which an observed change is statistically unlikely to reflect ordinary score fluctuation rather than true symptom change. A youth or parent may therefore notice a change that exceeds the MID before that change is large enough to exceed the RCI—a shift that feels different is not yet distinguishable from a good week versus a bad week until it also clears the RCI threshold.

Both are useful for MBC, because they answer different questions: MID flags when a change may be subjectively meaningful, and RCI flags when a change can be trusted as a reliable signal rather than measurement noise.

Present results also help clarify the complementary roles of parent- and youth-report measures. The PGBI-10M showed somewhat stronger internal consistency and, in ABCD, was the only instrument that could be evaluated against concurrent KSADS diagnoses because of Year alignment. The 7-Up nevertheless demonstrated useful psychometric performance in the same population context, supporting its role as a self-report companion for assessing activation and manic-spectrum symptoms. In practice, this strengthens the case for multi-informant assessment. Parent and youth reports need not converge perfectly to be clinically informative. Discrepancies may highlight the difference between externally visible behavioral dysregulation and internally experienced activation. Current findings support parallel use of the two scales for assessment, even though only the PGBI-10M could be tested here as a concurrent classifier.

The longitudinal DIF findings merit brief comment. For the PGBI-10M, 6 of 10 items showed threshold DIF from Year 0 to Year 2; for the 7-Up, nearly all non-anchor items showed threshold DIF from Year 1 to Year 3. Plausible contributors include developmental shifts in the behavioral baseline for concrete-referent items (e.g., activity level, talkativeness), maturing self- /caregiver differentiation of internal-state items (e.g., elevated mood), and period effects given the multi-year gap between waves. Consistent with this account, the PGBI-10M items that remained fully invariant across waves — sleep disruption, general mood pattern, and episodic rage — were the most behaviorally observable items and least dependent on caregiver inference about internal states, whereas items requiring more inferential judgment were more likely to show DIF. Disentangling these will require additional ABCD waves. The practical implication is that raw sum scores compared across waves conflate true change with item drift for non-invariant items. We recommend anchor-item or partial-invariance factor scores for tracking within-person change over time, reserving full-scale raw sums for cross-sectional use.

Several limitations should temper interpretation. First, ABCD data were from youths ∼9-12 years old; they should be extrapolated to other age-groups with caution, if at all. Second, KSADS-based bipolar diagnoses are stronger than unstructured community impressions, but no criterion is perfectly reliable, and criterion error constrains the maximum observable discrimination of any checklist. The ABCD KSADS is computer administered and uses an algorithm, not clinical judgment or a consensus review, to assign the diagnoses. The 2% rate of computerized KSADS bipolar diagnoses is at the high end of the range of plausible rates in the general population,^23,24^ and there are concerns that the computerized version has different validity than traditional KSADS diagnoses based on a trained interviewer.^41,42^ Yet even at a prevalence rate of 2% there is limited precision for subgroup and sensitivity analyses. The age range included here may attenuate discrimination because youth have not yet passed through the peak developmental window for onset of bipolar I or bipolar II disorder. As the classification analyses were concurrent, they do not answer the important question of whether early elevations on these scales forecast later bipolar outcomes. Bipolar disorder is partly heritable, meaning siblings might simultaneously experience elevated symptoms, and shared method variance may also be attributable to parents’ reporting on more than one participating child. ABCD is large and diverse but not identical to a nationally representative epidemiologic sample: generalize with caution. Also, the PGBI-10M and 7-Up in ABCD used a 12-month recall window, matching the annual assessment interval, but not well suited to detecting change attributable to recent clinical action. Shorter-window versions of these scale families have been used to track symptom change over shorter intervals in treatment contexts^17,18^. Against these limitations, the study’s strengths are substantial: a very large community cohort, parent- and youth-report mania measures, structured diagnostic data, and direct integration of psychometric benchmarking with clinically relevant classification analyses.

Future work should extend these analyses longitudinally, algorithmically. It also would be valuable to have more representative norms, more work with shorter retest intervals, and rating windows more consonant with MBC applications. Prior work suggests that family history may have a modest but incremental value in risk estimation^43^. Risk calculators have already shown promise for predicting bipolar outcomes in familial and clinical high-risk youth, and that line of work is a more plausible next step than refining a single universal screening threshold. A practical goal would be to combine repeated mania ratings with family history, depressive symptoms, anxiety, mood lability, functional change, and treatment context into individualized risk tools that support earlier identification without inflating false positives. At the health-system level, embedding such tools and ABCD-based change benchmarks into digital dashboards could make these measures more useful in routine care by helping clinicians and families distinguish signal from noise across repeated visits.

In sum, brief mania scales performed well psychometrically in a large population cohort, and the PGBI-10M retained statistically significant concurrent discrimination of KSADS-defined bipolar spectrum disorders. These findings point toward three complementary, indication-based uses for the PGBI-10M and 7-Up rather than a single blanket application. First, given the low positive predictive values reported above, the scales should not be used as universal screens in unselected populations. Second, they are well suited to targeted use when some other signal already raises concern—family history of bipolar disorder, depressive or mood-lability symptoms, treatment-emergent activation, or clinical impression—consistent with a Bayesian, evidence-based assessment approach. Third, the population-referenced benchmarks established here support a single-timepoint referral-triage use case distinct from serial monitoring: a provider encountering a youth with activation symptoms can use these thresholds to judge whether the symptom level is unusual enough, relative to age peers, to warrant referral for comprehensive evaluation, without requiring repeated administration or treating the scale score as a diagnostic verdict. Because this triage application relies on a single administration rather than on detecting change, it is not affected by the recall-window limitation described below. Existing MBC and treatment-response benchmarks for the PGBI-10M and 7-Up come from smaller clinical samples using shorter recall windows. Present findings complement, rather than replace, that work. Yet the same data also show the limits of universal screening for a low-base-rate condition: even a good measure will produce many false positives when applied indiscriminately. The practical lesson is not to abandon brief mania measures, but to use them where they are strongest: to support targeted, multi-informant assessment, to inform single-timepoint referral decisions using population-referenced benchmarks, and, where shorter-interval clinical benchmarks are available, to monitor clinically meaningful change in youths for whom mood activation is already a plausible concern.

## Data Availability

The data supporting the findings of this study are available from the corresponding author upon reasonable request.

## Author’s Contributions

EAY led the conceptualization. EAY, AJT, and PAR developed the methodology. EAY implemented the software. EAY, PAR, and YL conducted the validation. EAY, AJT, and YL performed the formal analysis. EAY, PAR, and DR provided the resources. EAY, AJT, and DR curated the data. EAY wrote the original draft. EAY, AJT, PAR, MBM, YL, DR, and CA reviewed and edited the manuscript. EAY and PAR created the visualizations. EAY supervised the project. EAY, AJT, MBM, YL, and DR managed the project administration. EAY completed the revisions and response to reviewers.

## Ethics Committee Approval

This study used secondary analyses of de-identified datasets. Each study received institutional review board approval at participating sites, and informed consent and assent were obtained from participants and their caregivers. The present analyses were conducted in accordance with local institutional policies and were considered exempt or not human subjects research.

---

**Supplemental Figure 1.**
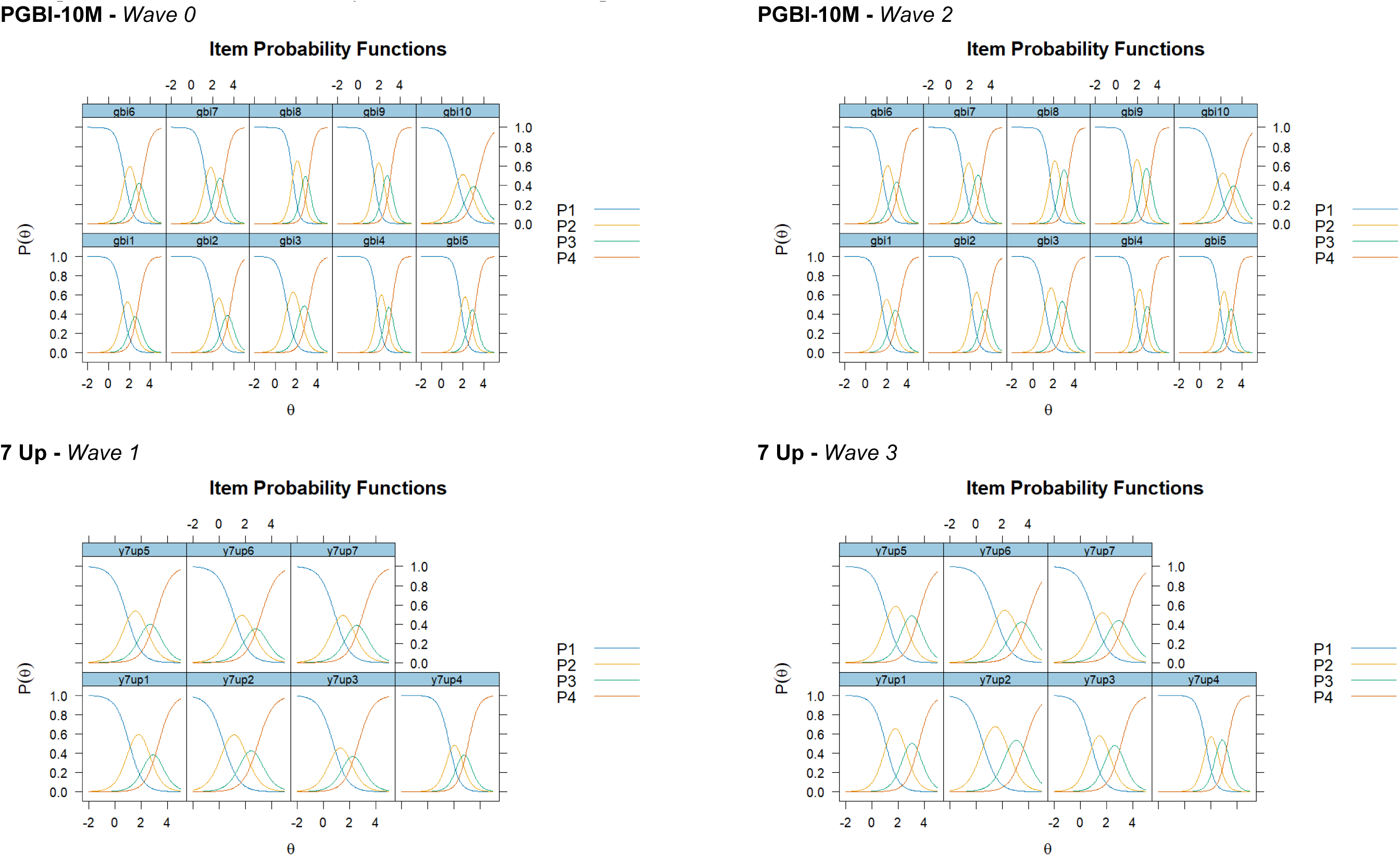
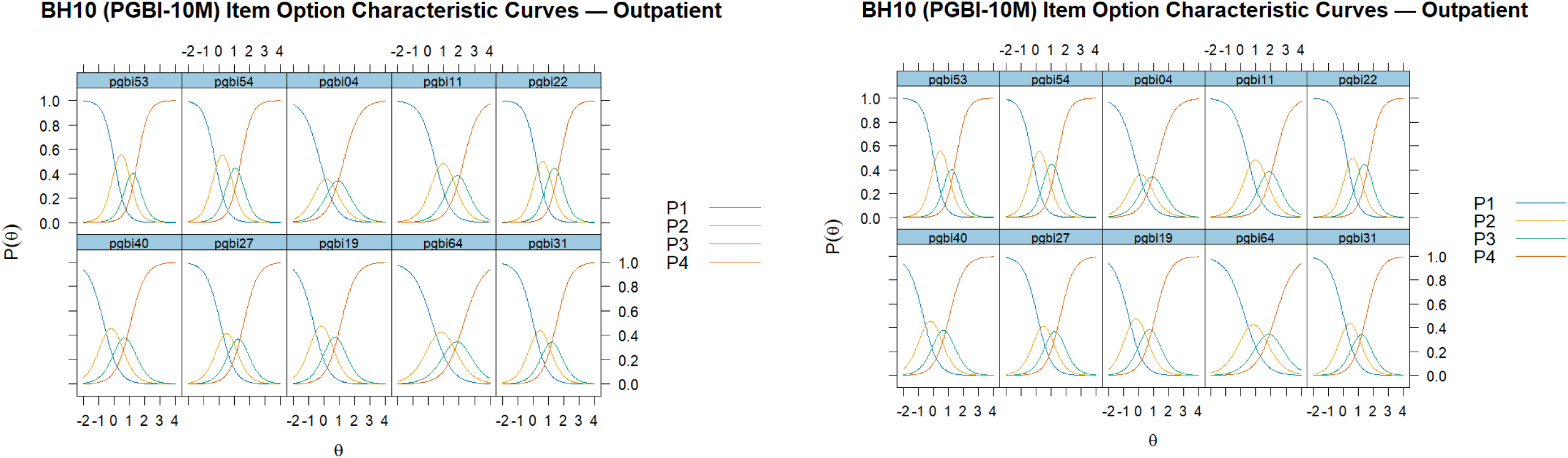
Item Option Characteristic Curves for PGBI-10M and 7-Up scales ***Notes.*** PGBI-10M = Parent General Behavior Inventory-10 Mania form. gbi6 = PGBI-10M’s item 6. y7up5 = 7-Up’s item 5. P1 = probability threshold 1 (transition from a response of 0 to a 1 on the item).

**Supplemental Table 1.** Item Parameters and Longitudinal Invariance Status, PGBI-10M (Wave 0 vs. Wave 2) Note. Slopes (a) are from the metric-invariance multigroup graded response model (equal across waves except items freed for slope DIF). DIF tests use Benjamini-Hochberg FDR-corrected p values; p < .05 is flagged as DIF. For items 1 and 9, the slope-DIF likelihood-ratio statistic was at or below zero (X^2^=-0.001 and 0.000, respectively), the strongest possible evidence of invariance rather than a missing test; a valid p value could not be computed from a non-positive chi-square statistic. Item 3 served as the anchor item (fixed slope and thresholds by design) and was not tested for threshold DIF. Shaded cells indicate items requiring freed parameters in the partial invariance model. See Supplemental Scale Items for full item text corresponding to items 1 to 10. Copies of the scales and scoring instructions are available at https://en.wikiversity.org/wiki/Evidence-based_assessment/Instruments/General_Behavior_Inventory

| Item | Original 73-item # | Item content | Anchor | Slope (Wave 0) | Slope (Wave 2) | Slope DIF (FDR p) | Slope Status | Threshold DIF (FDR p) | Threshold Status |
| --- | --- | --- | --- | --- | --- | --- | --- | --- | --- |
| 1 | 4 | Has your child experienced periods of several days or more when, although he/she was feeling unusually happy and intensely energetic (clearly more than your child's usual self), he/she was also physically restless, unable to sit still, and had to keep moving or jumping from one activity to another? | No | 2.5 | 2.5 | n.s. ($\chi^2=-0.001$) | Invariant (fixed) | 0.001 | DIF (freed) |
| 2 | 11 | Have there been periods of several days or more when your child's friends or other family members told you that your child seemed unusually happy or high – clearly different from his/her usual self or from a typical good mood? | No | 2.65 | 2.65 | 0.0283 | DIF (freed) | 0.0304 | DIF (freed) |
| 3 | 19 | Has your child's mood or energy shifted rapidly back and forth from happy to sad or high to low? | Yes | 2.46 | 2.46 | 0.0623 | Anchor (fixed) | — | Anchor (fixed) |
| 4 | 22 | Has your child had periods of extreme happiness and intense energy lasting several days or more when he/she also felt much more anxious or tense (jittery, nervous, uptight) than usual (other than related to the menstrual cycle)? | No | 3.36 | 3.36 | 0.1818 | Invariant (fixed) | 0.1782 | Invariant (fixed) |
| 5 | 27 | Have there been times of several days or more when, although your child was feeling unusually happy and intensely energetic (clearly more than his/her usual self), he/she also had to struggle very hard to control inner feelings of rage or an urge to smash or destroy things? | No | 3.28 | 3.28 | 0.0623 | Invariant (fixed) | 0.0184 | DIF (freed) |
| 6 | 31 | Has your child had periods of extreme happiness and intense energy (clearly more than his/her usual self) when, for several days or more, it took him/her over an hour to get to sleep at night? | No | 2.56 | 2.56 | 0.127 | Invariant (fixed) | 0.4086 | Invariant (fixed) |
| 7 | 40 | Have you found that your child's feelings or energy are generally up or down, but rarely in the middle? | No | 2.71 | 2.71 | 0.0623 | Invariant (fixed) | 0.0888 | Invariant (fixed) |
| 8 | 53 | Has your child had periods lasting several days or more when he/she felt depressed or irritable, and then other periods of several days or more when he/she felt extremely high, elated, and overflowing with energy? | No | 3.19 | 3.19 | 0.3293 | Invariant (fixed) | 0.0059 | DIF (freed) |
| 9 | 54 | Have there been periods when, although your child was feeling unusually happy and intensely energetic, almost everything got on his/her nerves and made him/her irritable or angry (other than related to the menstrual cycle)? | No | 3.17 | 3.17 | n.s. ($\chi^2=0.000$) | Invariant (fixed) | 0.0223 | DIF (freed) |
| 10 | 64 | Has your child had times when his/her thoughts and ideas came so fast that he/she couldn't get them all out, or they came so quickly others complained that they couldn't keep up with your child's ideas? | No | 1.84 | 1.84 | 0.127 | Invariant (fixed) | 0 | DIF (freed) |

**Supplemental Table 2.** Item Parameters and Longitudinal Invariance Status, 7-Up (Wave 1 vs. Wave 3) Note. Slopes (a) are from the metric-invariance multigroup graded response model. DIF tests use Benjamini-Hochberg FDR-corrected p values; p < .05 is flagged as DIF. y7up3 served as the anchor item (fixed slope and thresholds by design) and was not tested for threshold DIF. Shaded cells indicate items requiring freed parameters in the partial invariance model. See Supplemental Scale Items for full item text corresponding to y7up1–y7up7. Copies of the scales and scoring instructions are available at https://en.wikiversity.org/wiki/Evidence-based_assessment/Instruments/General_Behavior_Inventory

| Item | Original 73-item # | Item content | Anchor | Slope (Wave 1) | Slope (Wave 3) | Slope DIF (FDR p) | Slope Status | Threshold DIF (FDR p) | Threshold Status |
| --- | --- | --- | --- | --- | --- | --- | --- | --- | --- |
| 1 | 22 | Have you had periods of extreme happiness and intense energy lasting several days or more when you also felt much more anxious or tense (jittery, nervous, uptight) than usual (other than related to the menstrual cycle)? | No | 2 | 2 | 0.1714 | Invariant (fixed) | 0 | DIF (freed) |
| 2 | 30 | Have there been times lasting several days or more when you felt you must have lots of excitement, and you actually did a lot of new or different things? | No | 1.72 | 1.72 | 0.4905 | Invariant (fixed) | 0 | DIF (freed) |
| 3 | 31 | Have you had periods of extreme happiness and intense energy (clearly more than your usual self) when, for several days or more, it took you over an hour to get to sleep at night? | Yes | 1.88 | 1.88 | 0.0061 | Anchor (fixed) | — | Anchor (fixed) |
| 4 | 38 | Have you had periods of extreme happiness and high energy lasting several days or more when what you saw, heard, smelled, tasted, or touched seemed vivid or intense? | No | 2.7 | 2.7 | 0.0081 | DIF (freed) | 0 | DIF (freed) |
| 5 | 43 | Have there been periods of several days or more when your thinking was so clear and quick that it was much better than most other people's? | No | 1.87 | 1.87 | 0.2852 | Invariant (fixed) | 0 | DIF (freed) |
| 6 | 46 | Have there been times of several days or more when you felt that you were a very important person or that your abilities or talents were better than most other people's? | No | 1.68 | 1.68 | 0.4356 | Invariant (fixed) | 0 | DIF (freed) |
| 7 | 64 | Have you had times when your thoughts and ideas came so fast that you couldn't get them all out, or they came so quickly that others complained that they couldn't keep up with your ideas? | No | 1.7 | 1.7 | 0.4356 | Invariant (fixed) | 0.0116 | DIF (freed) |

